# Rising evidence of COVID-19 transmission potential to and between animals: do we need to be concerned?

**DOI:** 10.1101/2020.05.21.20109041

**Authors:** Andrei R. Akhmetzhanov, Natalie M. Linton, Hiroshi Nishiura

**Affiliations:** Graduate School of Medicine, Hokkaido University, Kita 15 Jo Nishi 7 Chome, Kita-ku, Sapporo-shi, Hokkaido 060-8638, Japan; Cluster Intervention Team, Ministry of Health, Labor, and Welfare, 1-2-2 Kasumigaseki Chiyoda-ku, Tokyo 100-8916, Japan; CREST, Japan Science and Technology Agency, Honcho 4-1-8, Kawaguchi, Saitama 332-0012, Japan

**Keywords:** COVID-19, domesticated animals, pets, transmission, basic reproduction number

## Abstract

Severe acute respiratory syndrome coronavirus 2 (SARS-CoV-2)—the virus that causes coronavirus disease (COVID-19)—has been detected in domestic dogs and cats, raising concerns of transmission from, to, or between these animals. There is currently no indication that feline- or canine-to-human transmission can occur, though there is rising evidence of the reverse. To explore the extent of animal-related transmission, we aggregated 17 case reports on confirmed SARS-CoV-2 infections in animals as of 15 May 2020. All but two animals fully recovered and had only mild respiratory or digestive symptoms. Using data from probable cat-to-cat transmission in Wuhan, China, we estimated the basic reproduction number *R*_0_ under this scenario at 1.09 (95% confidence interval: 1.05, 1.13). This value is much lower than the *R*_0_ reported for humans and close to one, indicating that the sustained transmission between cats is unlikely to occur. Our results support the view that the pet owners and other persons with COVID-19 in close contact with animals should be cautious of the way they interact with them.

## 1. Introduction

Severe acute respiratory syndrome coronavirus 2 (SARS-CoV-2)—the virus that causes coronavirus disease (COVID-19)—has been detected in domestic dogs and cats, raising concerns of transmission from, to, or between these animals. In late March 2020, a report from Hong Kong indicated two of nineteen dogs and one of nine cats from households with confirmed human COVID-19 cases or persons in close contact with confirmed patients tested positive for the virus [1,2], and in Belgium a cat in a household with a confirmed COVID-19 case was determined to be infected [3]. From April through the beginning of May, an additional six cases of COVID-19 among domestic cats were confirmed in Belgium, France, Spain, and United States [4]. While there is currently no indication that feline- or canine-to-human transmission can occur, there is some evidence of possible human-to-feline or human-to-canine transmission [2,5].

Animal-to-animal transmission also appears plausible. In a recently published preprint [6] on infections in cats in the original epicenter of COVID-19—Wuhan, China—researchers examined serological samples from domestic and stray cats samples collected during January and February 2020, among which 15% were positive. Some positive samples were from cats cared for by human COVID-19 cases or cats at a pet hospital, but samples from stray cats with no known link to human COVID-19 cases were also positive, indicating the possibility of sustained feline-to-feline transmission among stray cats.

Domestic animals are not the only ones affected. In early April, a 4-year-old female Malayan tiger cub tested positive for SARS-CoV-2 in New York, and six other tigers and lions at the zoo had symptoms consistent with the illness and were considered presumptive positive cases. The virus was likely transmitted to the cats from an asymptomatic zookeeper, though whether all the cats were infected by a human keeper or feline-to-feline transmission occurred after initial introduction of the virus to the cats is uncertain.

In addition, in late April the Dutch Ministry of Agriculture, Nature and Food Quality notified the World Organisation for Animal Health (OIE) of an outbreak of COVID-19 on four mink farms used for fur production. Three of the farms were epidemiologically linked to each other, but the fourth farm had no connection with the previous outbreaks. Two more infected farms were reported in early May. Although human-to-mink transmission is considered the probable source of the outbreaks, transmission between minks was indicated as the main driver [7]. In contrast to infected domestic dogs and cats, which are typically asymptomatic or have only a mild course of disease following infection, some infected minks suffered from a more severe course involving pneumonia and death [7].

Experimental evidence [8,9] has classified mustalids (mink and ferrets), felids (cats, both domestic and exotic), and rabbits as highly susceptible to COVID-19, while canids, domesticated birds and pigs are less likely to become infected and transmit the disease. In the experiments of [8], the authors inoculated infected animals with high viral loads. However, the majority of serological samples collected from stray and domestic cats showed a relatively weak viral titer [6]. Similarly, the viral titers of an infected dog in Hong Kong were also weak, and the dog was never considered contagious to other dogs or to humans [10]. In light of this information, rampant transmission of SARS-CoV-2 among stray cats in Wuhan seems unlikely. However, if sustained transmission among domesticated animals is possible, SARS-CoV-2 may pose a substantial threat to the animal communities themselves. There is also a risk that these animals will act as a reservoir for the virus [11].

In this paper, we attempt to resolve some of the uncertainty around SARS-CoV-2 transmission potential to and between animals and quantify transmission strength in mathematical terms. To do so, we 1) aggregated all publicly available records on confirmed cases of COVID-19 among animals, and 2) estimated the basic reproduction number *R*_0_—the average number of secondary cases generated by a single primary case—among cats using the results of the serological study of Zhang et al. [6] in our modeling framework. From this data and analysis we draw some inferences on transmissibility and discuss their implications for the current COVID-19 pandemic.

## 2. Methods

### 2.1. Epidemiological data

We retrieved information on confirmed SARS-CoV-2 animal cases from publicly available immediate notifications and follow-up reports in the World Animal Health Information System (WAHIS), published by the World Organization for Animal Health (OIE) (https://www.oie.int/wahis_2/public/wahid.php/Wahidhome/Home). The information on infected captive felids at the Bronx Zoo, New York, USA was additionally verified by the press releases of the Wildlife Conservation Society, a nonprofit organization which runs the Bronx Zoo (https://newsroom.wcs.org). In order to avoid missing possible reports, we also scanned the ProMED-mailing list (http://promedmail.org) using the query “covid AND animals.”

### 2.2. Basic reproduction number

In our modeling framework for the possible sustained chain of transmission among domestic and stray cats in Wuhan, China [6], we assumed homogeneous mixing and a random independent sampling process in the collection of the serological samples. We employed a formula on the final size of the epidemic and estimate the basic reproduction number *R*_0_. Given that *n* is the number of positive tests taken among total *N* samples, then the fraction of positives *z* in the entire population can be estimated by using the binomial likelihood:

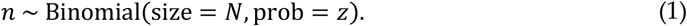

The basic reproduction number *R_0_* can be linked to the value of *z* as follows:

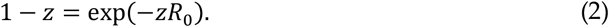

Knowing the value of *R_0_*, we also estimated the probability of major outbreak p according to the formula:

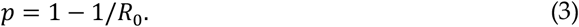

Its derivation relies on the assumption that infectious period follows an exponential distribution [12].

The statistical inference was done in a Bayesian framework by running Markov chain Monte Carlo (MCMC) simulations in Stan (cmdStan, version 2.23.0). The code is available at http://github.com/aakhmetz/Covid19-cats-story.

## 3. Results

Table 1 presents an aggregated list of 15 animal cases with confirmed SARS-CoV-2 infection. All cases recovered from the disease except two. In the first case, a dog from Hong Kong (D1)—the first reported case of COVID-19 in animals — died two days after release from quarantine, but its death was not linked to SARS-CoV-2 infection. In the second case, a cat from Spain (C4) was euthanatized in a veterinary clinic due to a severe heart problems. Three other animals from Hong Kong (D1–2 and C2) did not exhibit any clinical signs of the infection, while the six cat cases from Europe and the United States (C1, 3, and 5–8) showed only mild respiratory or digestive symptoms. All captive felids at the Bronx Zoo in New York (F1–7) exhibited a dry cough and some wheezing but did not shown any respiratory distress, though one animal lost their appetite.

**Table 1.**
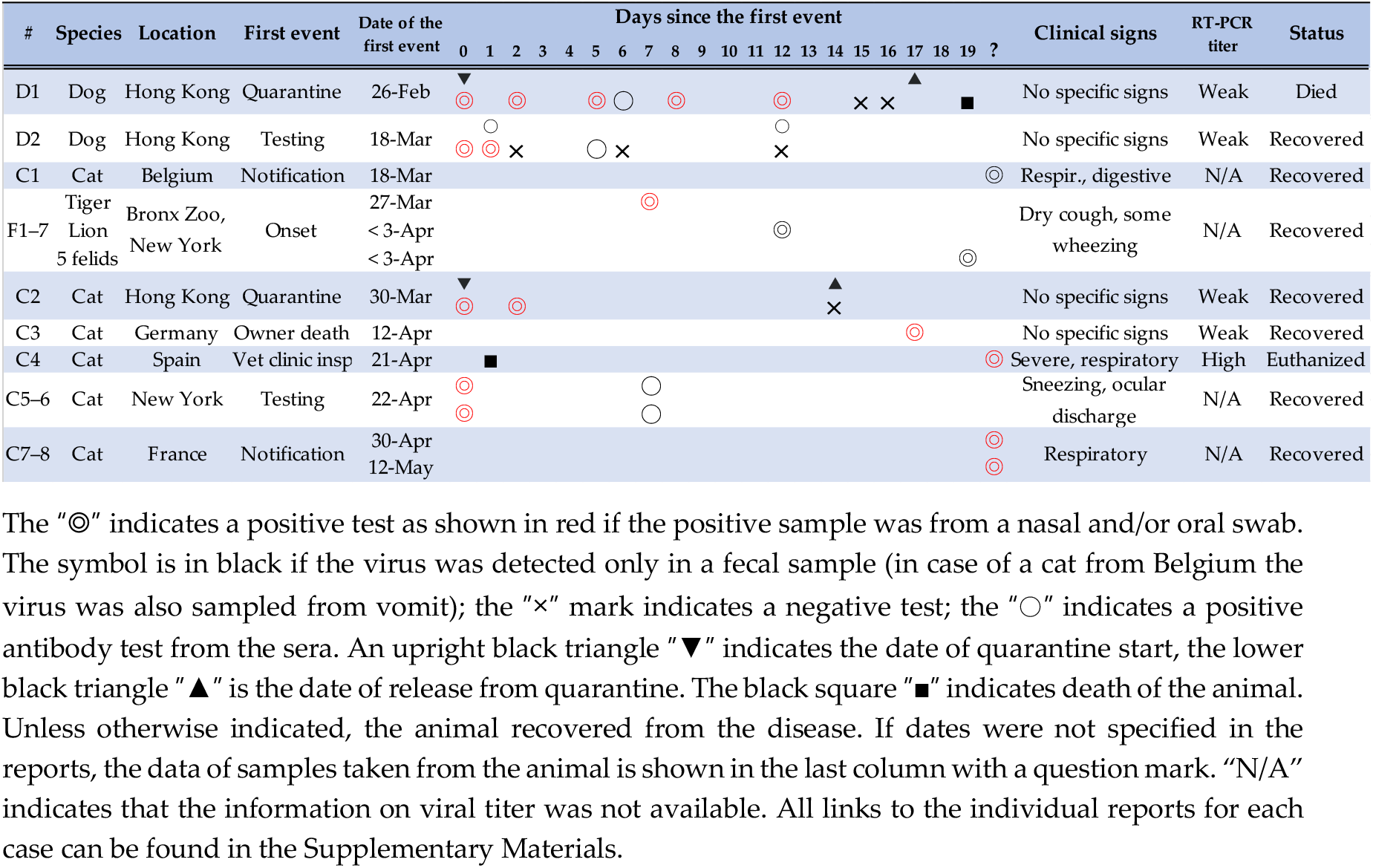
Confirmed COVID-19 cases among animals as of 15 May 2020

Of note, all cases among domestic animals except one were linked to the households with SARS-CoV-2 infected owners. The one unlinked case was a cat in New York (C3) who lived in a healthy household but was allowed to walk outside and likely was infected in the neighborhood.

Interested in whether transmission within cat populations could be occurring, we turned our attention to the study of Zhang et al [6] on serological samples from domestic and stray cats in Wuhan, China. Of serological samples collected during January and February 2020, 15% were positive. Some positive samples were from cats cared for by human COVID-19 cases or at a pet hospital, but stray cats with no known link to human COVID-19 cases tested positive as well. Following the modeling framework (1)–(2), we estimated the basic reproduction number *R_0_* at 1.09 (95% confidence interval [CI]: 1.05, 1.13), which suggested the probability of major outbreak at around 7.9% (95% CI: 4.7%, 12.1%). Given this low probability, the likelihood that sustained transmission among domestic and stray cats occurred in Wuhan, China is low.

## 4. Discussion

In Hong Kong, pets of human COVID-19 cases are generally quarantined and tested [13], which favors the detection of mild and asymptomatic cases. However, in other countries only symptomatic pets are likely to be detected typically no testing is done unless the animals are symptomatic. In line with this logic, all three cases in Hong Kong did not show any specific symptoms of COVID-19, while cases reported elsewhere exhibited respiratory or digestive symptoms. The observed difference in severity of disease for the animals may additionally be explained by the difference in prevalence of the COVID-19 human cases—as of 15 May, Hong Kong reported just over one thousand cases, while the other countries with domestic animal cases reported >50,000 human cases.

The secondary attack rate of SARS-CoV-2 from human to pets appears to be low and may also be driven by the infectiousness profile of their owners. The government of Hong Kong reported that about 50 animals including 2 hamsters were tested as of 17 April 2020, however, the count of confirmed cases remained the same compared to the earlier numbers [14]. A recent preprint [15] reported absence of SARS-CoV-2 infections among 9 cats and 12 dogs whose owners—veterinary students from the 20-year-old age group—formed a cluster of COVID-19 cases in France. Among 18 infected individuals, 2 were confirmed with PCR testing, 9 were presumptive cases, and 7 were asymptomatic [15]. On one hand, this may suggest that there was a lower infectiousness of 20-year-old individuals compared to pet owners with a more heterogenous age profile. On the other hand, the presence of asymptomatic human carriers could also play a role as some studies have shown highlighted the lack of secondary infections in households with asymptomatic index cases [16,17].

Historical records on the closely related virus SARS-CoV-1 reveal a similar story. During the 2003 outbreak of SARS, cats and ferrets were experimentally infected with the SARS-CoV-1 [18] but community transmission from humans to domestic animals was documented only once—in the Amoy Gardens complex in Hong Kong, where two blocks had a high level of contamination and eight cats and one dog tested positive from rectal swabs [19]. Transmission between animals and from animals to humans in the affected areas was only speculated [20]. In contrast, animals can be infected, even lethally, with seasonal influenza virus transmitted from their owner [21]. The human-to-animal transmission potential of SARS-CoV-2 likely lies somewhere between that of SARS-CoV-1 and influenza [22].

Our estimate of *R_0_ =* 1.09 for transmission between cats is much lower than the estimate of 2.4 for humans in the same area [23]. However, our results are subject to a few limitations. First, the data shared by [6] do not allow us to distinguish the negative serological tests of domestic cats from those of stray cats. We therefore assumed domestic cats to be part of the chain of transmission among all cats. We also assumed that stray cats were unlikely to be in close contact with humans. However, some stray cats could be fed by humans and there is a chance that the virus could be transmitted by an infected human to a stray cat by a contaminated feeder or other fomite. Second, we assumed that interventions implemented by the Chinese government to halt the epidemic in Wuhan in February-March 2020 did not affect the transmission dynamics of the virus among cats. Third, our calculation of the probability of a major outbreak—estimated at 7.9%—relied on the assumption of a homogeneous contact pattern between stray and domestic cats and imposed an exponentially distributed infectious period. The use of a heterogeneous contact pattern and (for example) a gamma distribution for the infectious period would provide more precise results [24–26], however incorporation of these factors requires more detailed data on the serological tests and the cat population in Wuhan than was available in the preprint by Zhang et al.

In conclusion, the current evidence supports the advice of veterinary authorities worldwide that the pet owners and other persons with COVID-19 in close contact with animals should pay special attention to the way they interact with them. There is no clear evidence that the virus is more than minimally transmissible to and among domestic animals, and the sporadic detections that are found are most likely to be only weakly positive. Therefore, most infections in domestic cats and dogs likely came directly from infected humans. At the same time, scientific evidence continues to indicate that the probability of SARS-CoV-2 transmission from domestic animals to humans is extremely low.

## Data Availability

All data sources are publicly available and do not require any approval.

## Author Contributions

A.R.A. conceived the study, collected and analyzed the data, A.R.A. and N.M.L. drafted the manuscript. All authors interpreted the data, edited the manuscript and approved the final version.

## Supplementary Materials

### List of sources used for Table 1

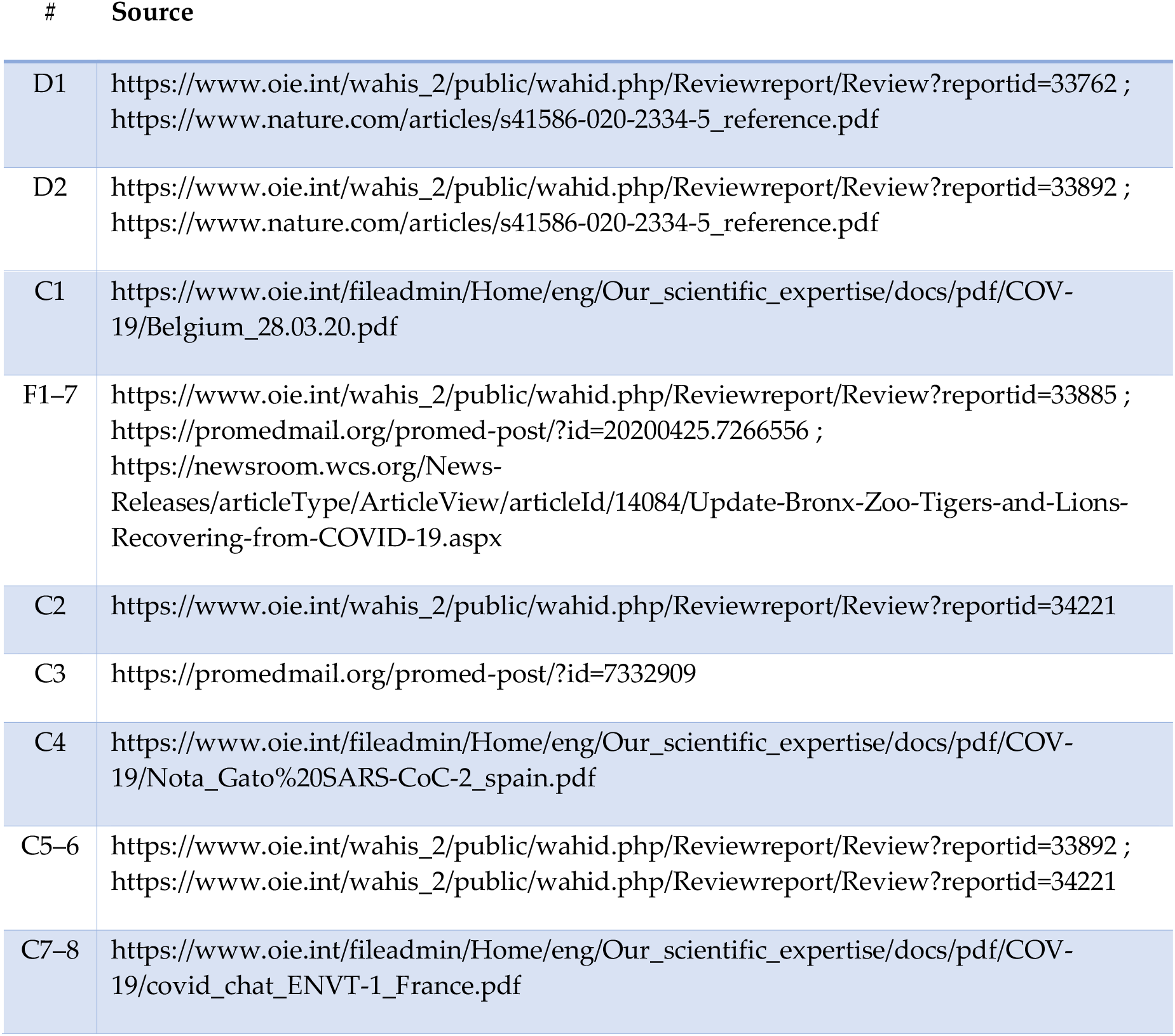

